# Assessing the Risk of Cascading COVID-19 Outbreaks from Prison-to-Prison Transfers

**DOI:** 10.1101/2021.04.12.21255363

**Authors:** Todd L. Parsons, Lee Worden

## Abstract

COVID-19 transmission has been widespread across the California prison system, and at least two of these outbreaks were caused by transfer of infected individuals between prisons. Risks of individual prison outbreaks due to introduction of the virus and of widespread transmission within prisons due to poor conditions have been documented. We examine the additional risk potentially posed by transfer between prisons that can lead to large-scale spread of outbreaks across the prison system if the rate of transfer is sufficiently high.

We estimated the threshold number of individuals transferred per prison per month to generate supercritical transmission between prisons, a condition that could lead to large-scale spread across the prison system. We obtained numerical estimates from a range of representative quantitative assumptions, and derived the percentage of transfers that must be performed with effective quarantine measures to prevent supercritical transmission given known rates of transfers occurring between California prisons.

Our mean estimate of the critical threshold rate of transfers was 14.38 individuals transferred per prison per month in the absence of quarantine measures. Available data documents transfers occurring at a rate of 60 transfers per prison per month. At that rate, estimates of the threshold rate of adherence to quarantine precautions had mean 76.03%. While the impact of vaccination and possible decarceration measures is unclear, we include estimates of the above quantities given reductions in the probability and extent of outbreaks.

We conclude that the risk of supercritical transmission between California prisons has been substantial, requiring quarantine protocols to be followed rigorously to manage this risk. The rate of outbreaks occurring in California prisons suggests that supercritical transmission may have occurred. We stress that the thresholds we estimate here do not define a safe level of transfers, even if supercritical transmission between prisons is avoided, since even low rates of transfer can cause very large outbreaks. We note that risks may persist after vaccination, due for example to variant strains, and in prison systems where widespread vaccination has not occurred. Decarceration remains urgently needed as a public health measure.

## 1. Introduction

As the COVID-19 pandemic continues in the United States, its dynamics in congregate settings of heightened transmission, including prisons, is crucial to understanding its spread, addressing racial disparities in the burden of the disease, and strategizing effective control.

Prisons are often overcrowded, unsanitary, and provide poor health care, and have been the site of many of the most concentrated and brutal outbreaks of the pandemic so far. Prevention of prison outbreaks is essential because standard control measures such as social distancing and self-isolation are not generally available to prison residents. One in five prisoners in the United States has been infected with SARS-CoV-2, compared to one in 20 in the U.S. overall, more than 1,700 have died, and prisoners continue to become infected.^1^ The New York Times reported on January 29, 2021 that of the ten largest outbreaks in U.S. correctional facilities to date, six of them have been in California state prisons.^2^ Every single one of California’s 35 prisons has reported 200 or more cases.^3^

Likely routes of introduction of the disease into prisons are via infected prison staffers, admission of infected prison residents from outside the prison system, and transfers of residents from other prisons. A widely reported outbreak at San Quentin prison in California, which infected over 2200 of the 3563 inmates and killed 28, was caused by a transfer of prisoners from the Correctional Institute for Men in Chino, California [16], and a subsequent outbreak at California Correctional Center in Susanville, California was likely caused by transfer from San Quentin. Multiple outbreaks in winter 2020 appear to have been caused by importation via staff members.

Prison outbreaks can be very large—the outbreak at San Quentin infected over 60% of the prison population, and the outbreak in California’s Avenal State Prison topped 80%—and because of well-known inequities in the criminal justice system, they contribute to racial inequity in the burden of COVID infection [9, 17, 13, 10]. In October 2020, the California Court of Appeals ruled that the California Department of Corrections and Rehabilitation (CDCR) has been guilty of “deliberate indifference” and that California prison populations must be reduced by half to address the ongoing risk of SARS-CoV-2 transmission.^4^ This decision reflects the recommendation to decarcerate California prisons to 50% of capacity published by public health experts during the San Quentin outbreak.^5^ The decision is undergoing appeal, substantial decarceration has not occurred, and multiple outbreaks have occurred in California state prisons in the time since the decision.

While the risks of outbreaks sparked by staff introductions or transfers and spread within prisons due to poor conditions are well known, here we examine the potential danger from another, potentially less apparent risk: the possibility that transmission from prison to prison via transfer of prison residents may be sufficient to lead to uncontrolled spread across the prison system. If such conditions should occur, the disease could be expected to spread to substantially more prisons than otherwise, and infect far more individuals (Figure 1). The risks to prison residents, staff, and surrounding communities could be considerably increased.

**Figure 1:**
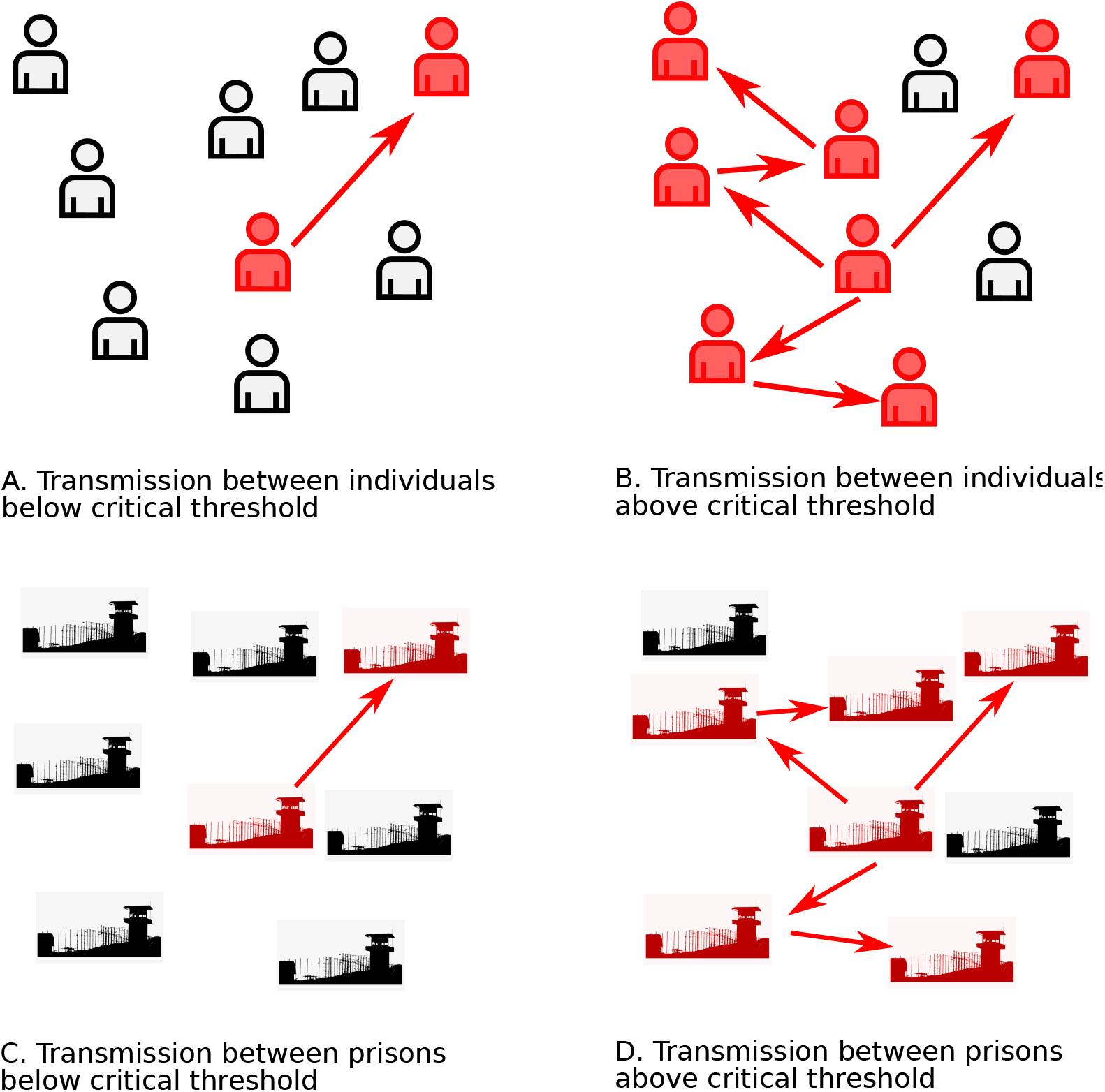
Scenarios for transmission between individuals and between prisons. A. When the reproduction number *R* between individuals — the mean number of cases caused by a case — is below the critical threshold of 1, transmission chains are short and outbreaks are small. B. When *R* is above the critical threshold, a large outbreak is possible. C. When the reproduction number *R*_*∗*_ between prisons — the mean number of prison outbreaks caused by a prison outbreak — is below the critical threshold of 1, transmission between prisons may still occur, but spread between prisons will be relatively limited. D. When *R*_*∗*_ is above the critical threshold, transmission between prisons can cascade and cause spread throughout the prison system.

The CDCR currently has quarantine and testing policies in place to prevent transfer of infective individuals. Unfortunately, adherence to CDCR policies has not always been universal, and it can not be assumed that no risky transfers occur.

We have addressed this question by using established theory of disease transmission, specifically a patch model to be defined below, to estimate the threshold rate of transfer associated with supercritical transmission between prisons, and the rate of adherence to transfer policies needed to prevent supercritical transmission at known rates of transfer.

## 2. Methods

A patch model of disease transmission can model a collection of discrete populations in which transmission happens within a population, and at a separate rate between populations. One such approach, the so-called *household model* [4], assumes that the population is divided into many small groups (the *households*) in which *local* contacts occur frequently, whereas *global* contacts may occur between any two individuals in the population, albeit at a much lower rate.

In such a model, there are two types of outbreaks: local ones, in which the infection spreads widely within a single group, but remains confined to that group, and global outbreaks, in which the epidemic spreads among many groups. Global outbreaks are governed by a group-to-group reproduction number *R*_*∗*_ whose value is the expected number of groups infected by transmission from a single group: the critical value is 1, and a large global outbreak is possible if the value is greater than 1 (Figure 1).

For our purposes, we assumed that all contacts are local, within groups, except when a transfer occurs of an individual from one group to another. We assumed also that the rate of transfer is low enough that an individual will transfer to at most one group while infective. We found (see Appendix) that the group-to-group reproduction number has the form

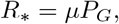

where *µ* is the total number of people infected in a group belonging to a randomly selected individual and *P*_*G*_ is the probability that an individual will transmit to a group other than the one where they were infected.

In order to evaluate how prison transfers affect prison-to-prison transmission, we modeled the group-to-group reproduction number in terms of the average number of individuals transferred between prisons per prison per month, a quantity which we call *n*. We expressed the above relation in terms of the transfer rate *n*, and solved for a threshold rate *n*^*∗*^ at which the critical value *R*_*∗*_ = 1.

We modeled an ensemble of scenarios for these quantities, to cover the range of possibilities.

1. **Optimistic vs. pessimistic reproduction number**. As an estimate of the basic reproduction number *R*_*L*_ within a prison we used the value 8.44 (95% credible interval: 5.00–13.13) estimated from a COVID-19 outbreak in a large urban jail in the U.S. [19]. Because this value is estimated from a setting in which a large outbreak occurred, and conditions in some prisons may be less conducive to transmission than those in which the largest outbreaks have occurred, we took the above number as a pessimistic estimate for *R*_*L*_. For an optimistic estimate, we calculated the probability *P*_*G*_ that a transfer event leads to transmission between prisons using the more optimistic value of 2.87 (95% CI, 2.39–3.44) that was estimated for a basic reproduction number for COVID-19 in general community transmission [5], and cut the probability in half to reflect the possibility that conditions may be better in roughly half of prisons.
2. **Optimistic vs. pessimistic attack rate within prisons**. Data from California Department of Corrections and Rehabilitation, collected by the UCLA Covid Behind Bars project,^6^ (Figure 2) provides outbreak sizes to date in California prisons. We took each prison at which the number of cases recorded to date is nonzero, which now includes all prisons in California, to represent an outbreak size to date. Using these numbers, the size-weighted mean outbreak size *µ* was 1513 cases. We took this value as a lower bound estimate of final outbreak sizes in California prisons. A conservative upper bound for *µ* may be the size-weighted mean of the overall population of each prison^7^ (see Appendix B), which was 2900.5 as of March 26, 2021.
3. **Optimistic vs. pessimistic secondary case distribution**. Evidence is accumulating that transmission of SARS-CoV-2 has an overdispersed pattern, in which many people cause few or no infections and a relatively large number of infections are caused by a few people [3, 2, 20]. This pattern may make the probability *P*_*G*_ lower than it could be, because relatively more people infect nobody. We estimated *P*_*G*_ given this pattern by assuming a negative binomial distribution of secondary cases, as is standard. However, this overdispersed pattern may be caused partially by wide variation in the number of people contacted by individuals socially [20], and it is not clear that this variation in contact structure is possible to the same degree in a prison setting, where individuals’ movements and locations are heavily constrained and regulated. For this reason, we also considered the possibility that secondary cases may be Poisson distributed within the prison setting, though they are more highly dispersed in community transmission.
4. **Optimistic vs. pessimistic timing of transmission events**. We also considered that the way in which the timing of transmission events is distributed can affect the probability of transmitting to another prison. If transmission tends to occur in bursts, for example driven by exceptional events when multiple people gather, such a burst might happen either before or after an individual is transferred from prison to prison. If transmission events are independent and happen all at different times, on the other hand, it is more likely that at least one of them will occur after a transfer. We model both of these cases.

We used each combination of the above assumptions to estimate a threshold transfer rate for the California prison system, above which transfers may create a risk of global spread of the coronavirus across the prison system. Calculations detailed in the Appendix provided a threshold value of the rate *ρ*_*G*_ of transfer between prisons per person per day for each combination of the above assumptions, which we transformed to a threshold rate *n*^*∗*^ of transfers per prison per month, using conversion factors of 30 days per month and the average number 2676.6 of individuals per prisons in the CDCR system as of March 26, 2021.

**Figure 2:**
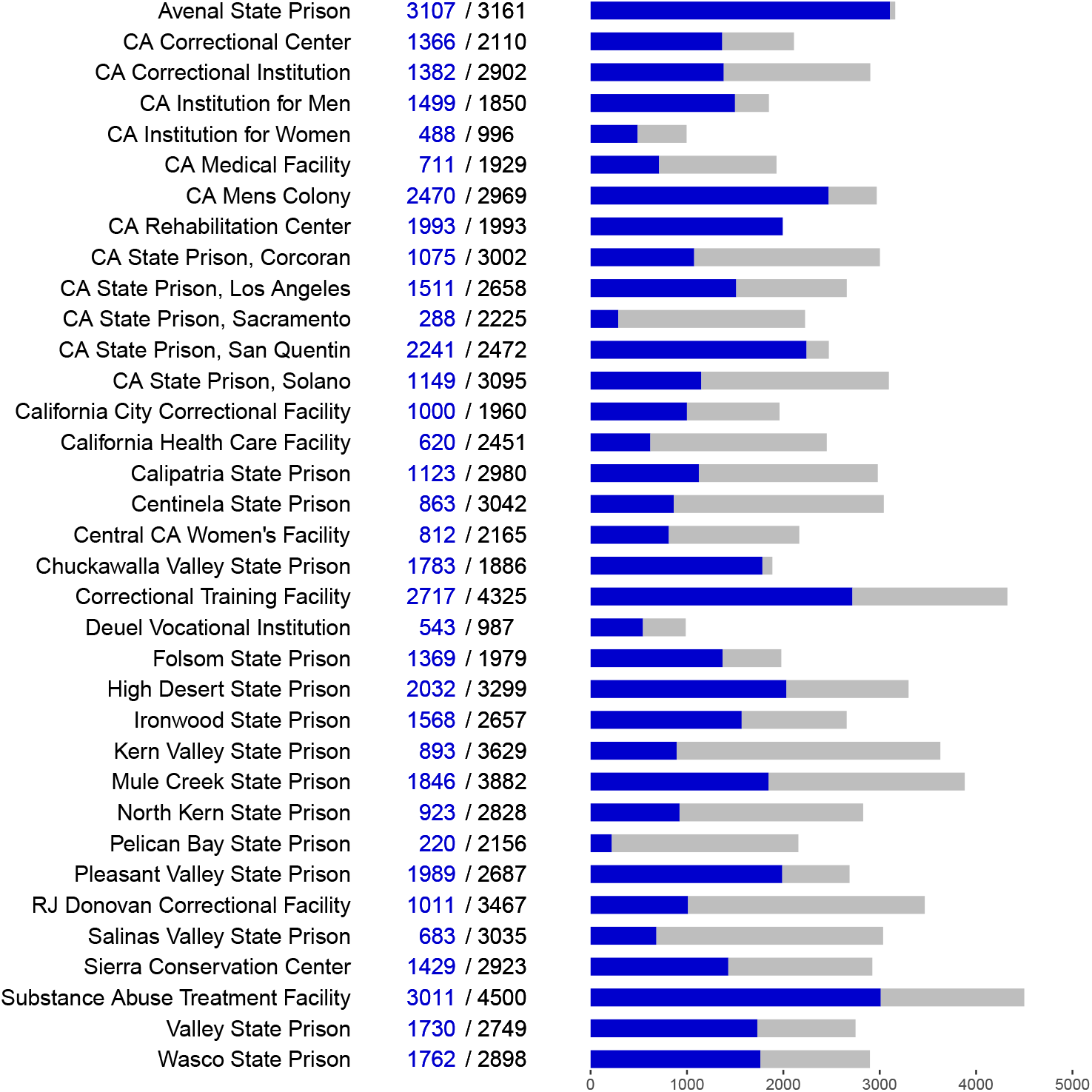
**Sizes of COVID-19 outbreaks in California prisons** as of March 26, 2021. Labels and bar charts show the number of cases to date (blue) and total population (black/gray) at each prison.

We note that this threshold number of transfers concerns potentially infective transfers who are exposed to the prison resident population in the facility where they arrive. Prisons, of course, have policies for quarantine of transferred residents and for testing before transfer to prevent transfer of infected individuals, and these policies are likely to reduce the risk due to transfer.

Transfers between California state prisons are regulated by a policy called the movement matrix^8^. Residents are tested five days before transfer, rapid tested one day before transfer, and quarantined for 14 days after transfer. Quarantine is in celled housing with a solid door if possible, and otherwise in cohorts of no more than four people. Residents in quarantine are screened for symptoms daily, tested if symptomatic, and isolated if they test positive. All of them are tested after five days post-transfer, again after 12 to 14 days post-transfer, and then released if negative and asymptomatic. Both residents and staff are to wear N95 masks during transfer. Residents who have been diagnosed with COVID-19 and subsequently resolved are considered immune for 90 days and exempt from quarantine and testing. After 90 days they are considered susceptible again and subject to the above measures.

We estimated that these procedures are likely to reduce the risk of transmission by transfer substantially. However, compliance with safety policies may not be perfect. For example, the California Inspector General has documented extensive noncompliance with mask guidelines in the California prisons, including during the San Quentin outbreak [15]. If protective policies reduce the number of transfers who can potentially transmit the virus by some percentage, it is the number of unprotected transfers that must be compared to the threshold value. If transfers are occurring at a known rate *n*_*t*_, and the threshold transfer rate for uncontrolled transmission between prisons is *n*^*∗*^, then the percentage of transfers that must be conducted in adherence to the protective policy in order to reduce unprotected transfers to the threshold rate is *a* = 100(*n*_*t*_ − *n*^*∗*^)*/n*_*t*_, or zero if *n*^*∗*^ exceeds *n*_*t*_.

### 2.1. Vaccination and decarceration

Vaccination in the California prison system is underway, with CDCR reporting that 40% of prison residents have received COVID-19 vaccination. A recent legal filing reported that accounting for previously infected prisoners, 76% of incarcerated people may have immunity [14].

Both increasing immunity and decarceration are likely to affect the spread of infections in at least two important ways, firstly by reducing the reproduction number and relatedly the probability that an introduction leads to an outbreak, and second by reducing the sizes of outbreaks if they occur. Both of these changes will affect our estimates of the rate of transfers needed to produce cascading outbreaks.

We look at the relation between increasing immunity and the critical threshold for cascading outbreaks by estimating the threshold transfer rate and associated quarantine adherence rate, as above, while reducing the local reproduction number parameters discussed above (*R*_*L*_) by half (which affects our estimate of how often a transfer causes an outbreak, but not of outbreak size), reducing the characteristic outbreak size *µ* by half, and reducing both by half simultaneously.

## 3. Results

We first estimated the threshold transfer rate (*n*^*∗*^) under all combinations of the above listed model assumptions, without protective measures (Table 1, Figure 3). The values estimated for *n*^*∗*^ ranged from 3.6 to 40.28 individuals transferred per prison per month, with mean 14.38. The generation time distribution used in these estimates was that estimated in a recent meta-analysis [8]: a Weibull distribution with mean 5.5 days and standard deviation 1.8 days (parameters *α* = 3.89, *β* = 6.08).

**Table 1:** Estimates of critical threshold for supercritical transmission between prisons in individuals transferred per prison per month and needed levels of adherence, given partial adherence with California’s transfer policy.

| Optimistic $\mu$ | Optimistic $R$ | Optimistic Case Distribution | Optimistic Timing | $n^*$ | Threshold Adherence |
| --- | --- | --- | --- | --- | --- |
| Y | Y | Y | Y | 40.28 | 32.87 |
| N | Y | Y | Y | 20.22 | 66.30 |
| Y | N | Y | Y | 14.20 | 76.33 |
| N | N | Y | Y | 6.18 | 89.70 |
| Y | Y | N | Y | 20.22 | 66.30 |
| N | Y | N | Y | 10.19 | 83.02 |
| Y | N | N | Y | 10.19 | 83.02 |
| N | N | N | Y | 4.17 | 93.05 |
| Y | Y | Y | N | 33.47 | 44.22 |
| N | Y | Y | N | 17.54 | 70.77 |
| Y | N | Y | N | 9.57 | 84.05 |
| N | N | Y | N | 5.59 | 90.69 |
| Y | Y | N | N | 17.54 | 70.77 |
| N | Y | N | N | 9.57 | 84.05 |
| Y | N | N | N | 7.58 | 87.37 |
| N | N | N | N | 3.60 | 94.00 |

**Figure 3:**
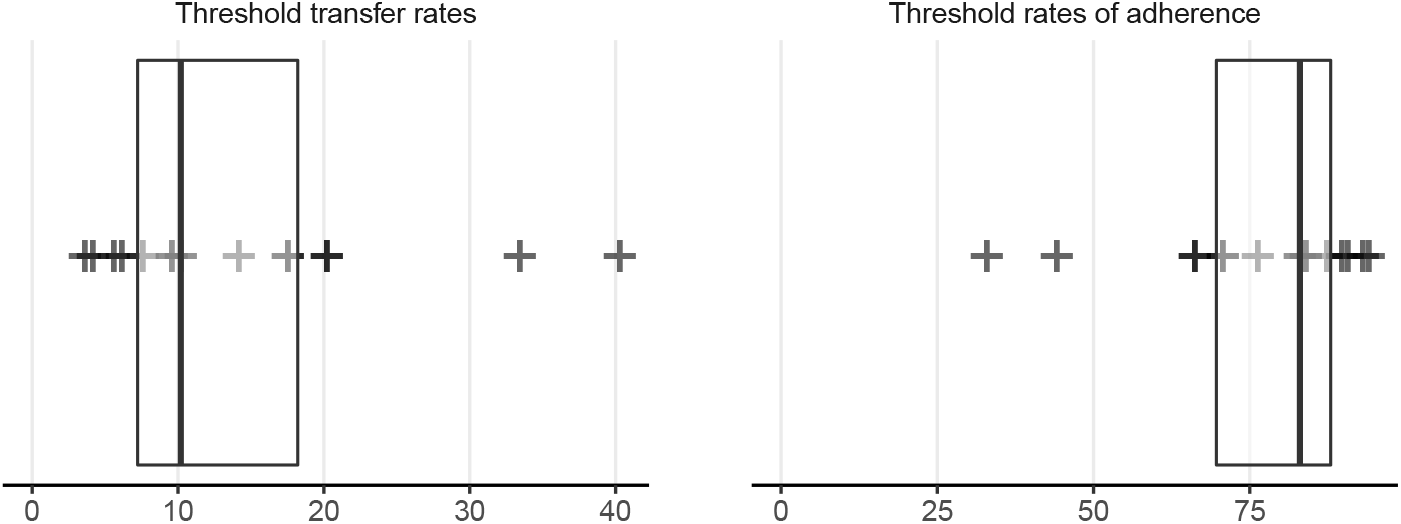
Estimated threshold transfer rates and levels of adherence to policy. (Left) threshold number of individuals transferred per prison per month, under multiple scenarios (Table 1), (Right) rate of adherence to transfer policy needed to reach threshold number of transfers, under the assumption of 60 total transfers per prison per month. Box plots display median and inter-quartile range.

Records released in the course of ongoing legal proceedings document the rate of transfers between prisons in the period from September 21 through October 11, 2020 at about 500 individuals transferred per week [1, p. 7, line 8]. There are 35 institutions in the California prison system, making that equivalent to about 60 transfers per prison per month. We converted our threshold estimates to the percentage of transfers that must be conducted in compliance with the safety policy in order to achieve the threshold number of unprotected transfers or below, given a total of 60 transfers per prison per month (Table 1, Figure 3). Our estimates of this threshold rate of adherence to quarantine precautions ranged widely but clustered in the upper third of the percentage scale, with mean 76.03%.

As a look at the sensitivity of our estimates to reductions in risk due to vaccination and/or decarceration, we estimated the same quantities while reducing the probability of an outbreak, the size of outbreaks, or both, by half (Figure 4). We estimated that while under our most optimistic assumptions the risk of cascading outbreaks is reduced substantially at 60 transfers per facility per month, to the point where quarantine measures could be ignored entirely without exceeding the estimated threshold (which could of course cause substantial risks other than cascading outbreaks), the change in the median and mean estimates is much more modest. We estimated the mean threshold transfers per prison per month (*n*^*∗*^) at 28.25, 28.88, and 57.11 respectively when reducing the assumed parameter *R* by half for estimation of the probability of an outbreak, reducing the characteristic outbreak size *µ* by half, and both. The mean estimate of threshold rate of adherence to policy was 59.35%, 54.76%, and 40.13% respectively.

**Figure 4:**
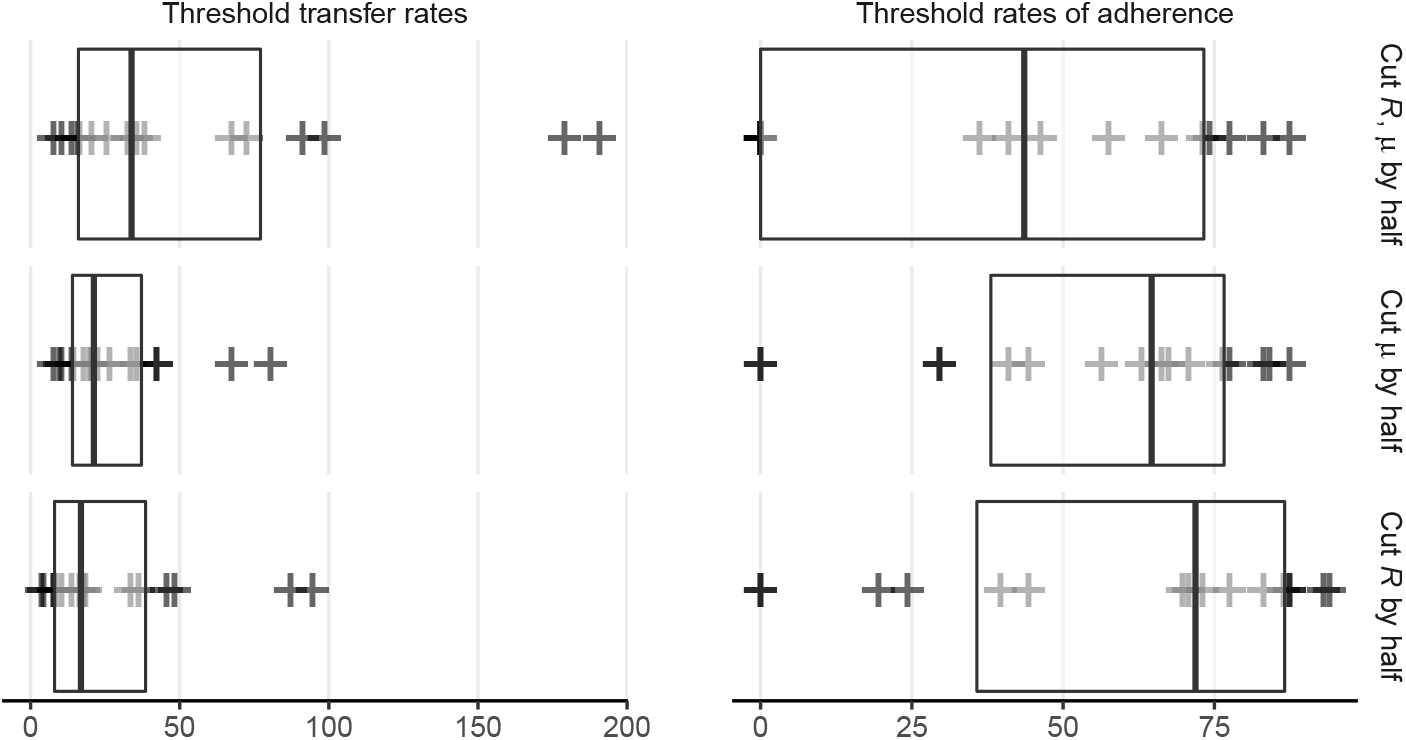
**Estimated threshold transfer rates and levels of adherence to policy given vaccination and/or decarceration**, as in previous figure with reductions in reproduction number and/or outbreak size assumed. (Left) threshold number of individuals transferred per prison per month, under multiple scenarios as above, (Right) rate of adherence to transfer policy needed to reach threshold number of transfers, under the assumption of 60 total transfers per prison per month. Box plots display median and inter-quartile range.

## 4. Discussion

We have constructed a range of values of a threshold rate of mixing between prisons above which transmission between prisons is likely to be supercritical. Supercriticality between prisons means that a prison outbreak is expected to produce more than one other prison outbreak on average, potentially leading to uncontrolled spread throughout the prison system.

We estimate that the reported rate of transfers that has been occurring in the California prison system has likely exceeded this threshold. We estimate that at these rates of transfers, the quarantine precautions must be highly effective and rates of compliance must be high to avoid risk of supercritical transmission between prisons. The rate of outbreaks occurring in California prisons suggests that supercritical transmission may already have occurred or may be occurring.

We offer these estimates as a way of assessing one of the multiple risks posed by infectous disease transmission in the prison system. It is important to note that this threshold can not be understood as providing a safe or acceptable rate of transfers, since transfer rates below the critical threshold can still cause huge outbreaks in multiple prisons. We are discussing an additional risk beyond the clear dangers of prisons’ unsafe conditions and spread due to transfers: the risk that in addition to having multiple large and deadly prison outbreaks of COVID-19, the rate of transfers could be sufficient to cause uncontrolled spread across a large portion of the prison system. This situation would likely lead to a great deal more harm to prison residents and staff than even the known risks of multiple large prison outbreaks, and could place communities throughout the state at risk as well.

The program of vaccination that is underway in the California prison system is crucial in reducing transmission and saving lives, and will likely help to end the pandemic more broadly as prison transmission poses risks to communities beyond the prison walls. We caution that substantial risks may continue to exist in the CDCR system, as spread of the SARS-CoV-2 virus can still occur, prison conditions continue to be overcrowded and unsanitary, and the effects of variant strains are yet unknown, not to mention the other diseases currently circulating and the potential of future emerging pandemics. Our results and methods may also be relevant to other prison systems where vaccination is not yet widespread. Decarceration remains a crucial public health measure to bring disease spread under control.

While these estimates are necessarily imprecise due to limited availability of data, such that risks could in fact be lower than we have estimated, we note as well that in addition to the mechanism of transfer of prison residents considered here, transmission between prison facilities may also be occurring resulting from travel of infected staff who work at multiple facilities. For this reason, the risk of uncontrolled transmission between prisons may in fact have been higher than we have estimated here.

These results have a number of limitations. We have assumed that individuals are removed from the epidemic process at the end of their infective period, as we consider the final size of each local epidemic, and thus do not account for the possibility of reinfection. In using a branching process, we have implicitly assumed a very large number of local communities, so that at least initially, each global transmission is to a new site, and ignores the possibility of a second epidemic in the same location. This assumption is reasonable in the context of prisons, where there are indeed many sites. The assumption of a homogeneous rate of transfer per individual across all prisons may be limiting as heterogeneity may be important.

This approach is applicable to analysis of risk due to transmission between sites in a variety of hotspot settings of transmission including but not limited to prisons. Transfer, migration, and mixing between sites may be important sources of risk in other locations of high transmission as well, such as jails, ICE facilities, skilled nursing care facilities, meat packing plants, and other agricultural operations.

## Data Availability

All data used in this manuscript is publicly accessible from sources listed in the manuscript.

## 5. Acknowledgements

LW acknowledges NIH R01 GM130900/GM/NIGMS. We are grateful to Travis Porco, Seth Blumberg, and Jianda Monique, to Martha Lincoln, Sarah Ackley, and multiple colleagues in the Amend and CalPROTECT projects and at CCHCS and CDCR for helpful comments on this work, and to Michael Bien and Ernest Galvan at Rosen, Bien, Galvan & Grunfeld LLP for helpful information regarding transfers.

## Appendix A. Patch Model Results

A patch model of disease transmission can model a collection of discrete populations in which transmission happens within a population, and at a separate rate between populations. One such approach, the so-called *household model* [4], assumes that the population is divided into many small groups (the *households*) in which *local* contacts occur frequently, whereas *global* contacts may occur between any two individuals in the population, albeit at a much lower rate. In such a model, there are two types of outbreaks, local ones, in which the infection spreads widely within a single subpopulation, but remains confined to that subpopulation, and global outbreaks, in which the epidemic spreads among many subpopulations.

A branching process approximation allows us to compute the probability that a global outbreak occurs: if there are a large number of households, the probability that two or more members of the same household are infected by individuals from different households is negligible, so, at the beginning at least, we can assume that each global contact is with a new household. Now, a branching process either goes extinct rapidly say with probability *q*, or grows indefinitely, with probability 1 − *q*. The latter corresponds to a major epidemic. Now we focus on the first individual infected, and the number that they infect in their household. If the number of households is large, then to first approximation, each of those individuals starts a branching process of infected households. If the branching process goes extinct, then necessarily, the branching processes started by each infected individual in the first household go extinct as well, which occurs independently for each branching process with probability *q* as well; this gives us a recursive formula for *q*.

Suppose that the fraction of households with *i* individuals is *h*_*i*_. If we were to choose a household at random, *h*_*i*_ gives the probability of choosing a household of size *i*; if, on the other hand we choose an *individual* at random, then the probability that individual comes from a household of size *i*, say *π*_*i*_, is proportional to *ih*_*i*_ (we have equal chance of choosing each individual from the same household). Now, given a household of size *i*, let *P*_*ij*_ be the probability that *j* ≤ *i* individuals in that household are ultimately infected. We then have

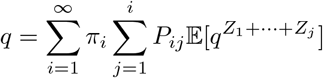

where *Z*_1_, …, *Z*_*j*_ are the number of *households* infected by each of the *j* infected individuals in the first household. We will assume that *Z*_2_, …, *Z*_*j*_ are identical and independent copies of a random variable *Z*, and all are independent of the *Z*_1_, the number of subsequent households infected by the first infected individual. Because the first individual has already infected one household, we have additional information on that individual, and *Z*_1_ must be distributed differently in light of that information; for example, the first individual may be symptomatic and thus will not be transferred again during their infectious period.

Thus,

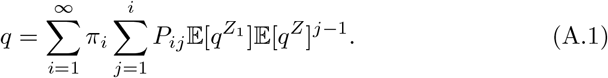

(*N.B*. it is here that we are assuming the number of households is very large, so that each infected individual is making contact with distinct households). Write *F* (*q*) for the expression on the right in (A.1); solving *q* = *F* (*q*) exactly is generally impossible, though we always have *F* (1) = 1. An epidemic is possible if and only if this equation has a second solution 0 *< q <* 1. Notice that

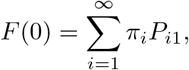

the probability that the first individual infects no other individuals in their household, so *F* (0) > 0. Now, a quick sketch shows that there is a second solution if and only if *R*_*∗*_ = *F*′(1) > 1:

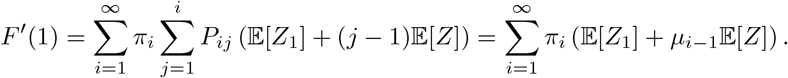

where *µ*_*i−*1_ is the average number of the *i* − 1 remaining individuals in a household of size *i* who are infected by the first infected (see Appendix B for an approximation of 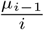 when *i* is large). Thus

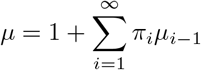

is the *size-biased* mean size of a local outbreak and

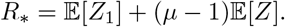

*R*_*∗*_ thus reflects the fact that transmission between households is proportional to the number of individuals within the house, who each individually make global contacts with other households. In the same way that the critical threshold *R*_0_ = 1 for the basic reproduction number separates subcritical from supercritical transmission in non-patch models, the boundary *R*_*∗*_ = 1 is the global critical threshold above which transmission is globally supercritical, meaning that an outbreak in one group is expected to infect more than one other group on average and can cause a large outbreak across the system of groups.

The branching process approximation we have chosen has considerable flexibility beyond the application presented here. At the cost of making the parameter *µ* something of a black box, we need only make minimal assumptions about individual epidemic dynamics: for example, we do not need to make specific assumptions about the duration of the infectious or latent periods, and the results are equally compatible with SIR or SEIR dynamics. Each of these choices, however, will result in different values of *µ*; while the distribution of prison sizes *π*_*i*_ is an empirical and observable quantity, the mean number of infections in a site will have to be computed. When individual facilities are sufficiently large that an ordinary differential equation (ODE) model is reasonable approximation to the local dynamics, we can use standard compartmental models of estimate the final size of the epidemic (see Appendix B for estimates for the SIR and SEIR models; unfortunately, adding a class of asymptomatic infectives, which is appropriate for COVID-19, leaves us unable to obtain exact results). When local facilities are small, more computationally intensive methods are required (see [18]).

We next turn our attention to computing 𝔼 [*Z*_1_] and 𝔼 [*Z*] in the context of transfer between prisons.

### Appendix A.1. Computing 𝔼[*Z*_1_] *and* 𝔼[*Z*]

Recall that, by assumption, an individual can infect a new site if and only if they are transferred during their generation interval. We will assume that transfers occur as a Poisson point process with rate *ρ*_*G*_, which for simplicity is sufficiently small that the probability that a given individual is transferred multiple times is negligibly small. Thus, each individual waits an exponentially distributed time with mean 1*/ρ*_*G*_ before being transferred. Under these assumptions, 𝔼[*Z*_1_] and 𝔼[*Z*] are simply the probability that the first infected individual or a subsequently infected individual, respectively, causes at least one new infection in the new site, say 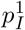 and *p*_*I*_ respectively.

To obtain a range of plausible values for *p*_*I*_, we will consider two extreme scenarios that bookend the degree of correlation between infection times:

i. A compound Poisson model in which infections happen individually and independently with a time-dependent rate, its *hazard function*, and
ii. A “burst” model in which all infections occur simultaneously; note that in this case, the first infected individual has already burst, so 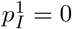.

We first present the former, and then show how the latter can be understood as a limiting case of a generalized version of the former.

#### Appendix A.1.1. The Hazard Function of the Infection Point Process and the Generation Interval Density

To derive the probabilities 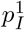 and *p*_*I*_, we first relate the generation interval density, *g*(*t*), to the hazard function of an inhomogeneous Poisson process. Fix *t* = 0 as the time that a given primary individual is infected, and assume secondary infections at random times *t*_1_, *t*_2_, If no two points coincide, *i.e*. if the infection is a *simple point process*, then it can be represented as an an inhomogeneous Poisson process (*n.b*., while no two points coincide, we do allow points to be arbitrarily close). It is convenient to represent the latter via its *counting measure, N* : given a subset *A* ⊆ [0, ∞),

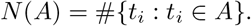

Because of our assumption of distinct points,

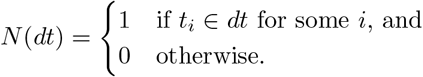

One can integrate with respect to *N* (*dt*):

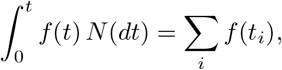

whenever the sum on the right hand side exists.

We define the *hazard function* of the point process, *h*(*t*) by

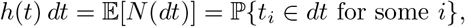

*i.e. h*(*t*) is the probability rate function for points. If 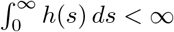, then we can normalize *h*(*t*) to obtain a probability density function,

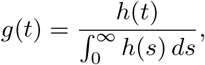

which is the generation interval density.

By definition of the hazard function,

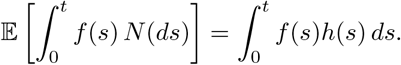

In particular, the total expected number of infections, the *local reproduction number*, is

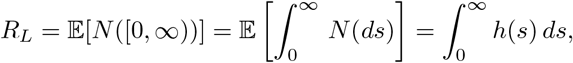

Thus, *h*(*t*) = *R*_*L*_*g*(*t*).

Now, by definition of the inhomogeneous Poisson process and its hazard function,

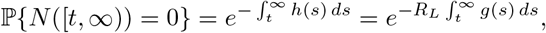

and, if we assume that transfer happens as a Poisson point process with rate *ρ*_*G*_, then the probability of no infections after transfer (1 − *p*_*I*_) is

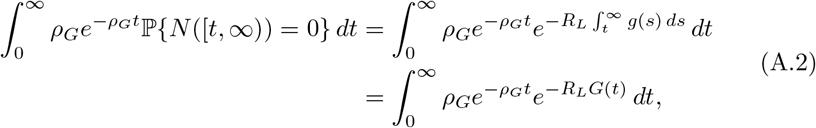

where we set 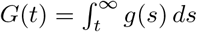. We can compute this integral numerically using a suitable choice of *g*(*t*), *e.g*. the empirical generation interval distribution.

For the first individual, we must account for the fact that they have already infected at least one other individual before transfer, and that, if symptomatic, we assume that the probability of transfer is greatly reduced, by some factor *f <* 1. Thus, rather than take time *t* = 0 to be the time of infection, we take it to be the time *T* of some (arbitrary) infection in the first site, so *T* is distributed according to *g*(*t*):

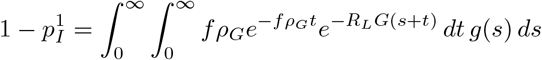

#### Appendix A.1.2. Compound Poisson Processes

More generally, we can consider the case when events occur according to a hazard rate *h*(*t*), but now the number of infections occurring at the *i*^th^ contact is given by i.i.d. random variables *ν*_*i*_; for example, if we can only poorly resolve infection times, we may be unable to separate them in time. We then have that the number of infections occuring at times in the set *A* is

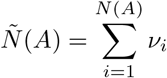

Then, as before,

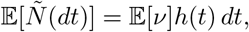

and

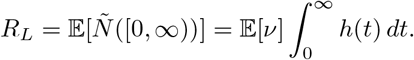

Unlike previously, this relation does not fix 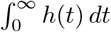: rather, for any *λ* ∈ (0, ∞), we can have 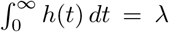, provided 𝔼[*ν*] = *R*_*L*_*/λ*. We then have *h*(*t*) = *λg*(*t*), so *λ* determines the intensity of points, whereas an increased intensity must correspond to fewer infections on average per event and *vice versa*. In particular, for a given *R*_*L*_, we can potentially have arbitrarily large clusters of infections, provided they occur sufficiently rarely.

#### Appendix A.1.3. Doubly Stochastic Poisson Processes and Overdispersion

Whilst the compound Poisson process allows for temporal clustering of infections, the number of infections in any interval [*a, b*] is Poisson distributed with rate 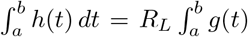 *dt* and thus has index of dispersion of 1. To allow for varying dispersion – as well as individual variation in contact rates – we can instead consider a doubly stochastic Poisson (or Cox) process, drawing an individual reproductive ratio, *r*, for each individual from a fixed distribution with mean *R*_*L*_.

If we assume each individual’s *r* is gamma distributed with shape parameter *k* and scale parameter 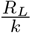 for some *k >* 0, then the probability of *m* infections in an interval [*a, b*] caused by a randomly chosen individual is

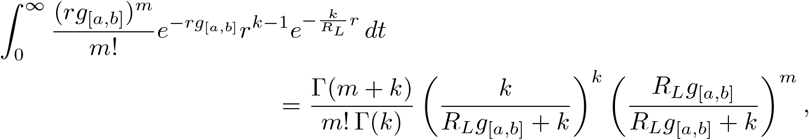

which we recognize as a negative binomial distribution with success probability 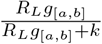 and *k* failures. Thus, the mean number of infections in [*a, b*] is thus *R*_*L*_*g*_[*a,b*]_ and the index of dispersion of infections in [*a, b*] is 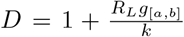. In particular, taking [*a, b*] to be the whole real line, we see that the mean total number of infections caused by a single individual is negative binomially distributed with mean *R*_*L*_ and index of dispersion 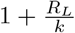.

Under this Cox process model, the probability of causing an infection after transfer is obtained by averaging (A.2) over a Gamma 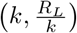-distribution. Because the reproductive ratio only appears in the exponential *e*^*−rG*(*t*)^, we can use the known probability generating function for the gamma distribution to get that

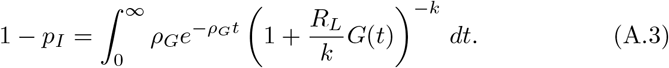

and

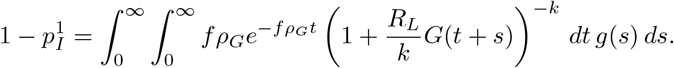

#### Appendix A.1.4. Conditioning on a Single Event

As an extreme case, we consider a scenario in which all transmission from a given individual occurs in a single event, for instance reflecting temporal heterogeneity in the number of social contacts. Observe that the probability of exactly one event in an inhomogeneous Poisson point process is

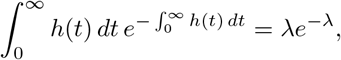

whereas the joint probability of having a single point at *t* is

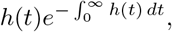

so the probability of a single point at *t* conditional on only one point is the ratio of these two probabilities,

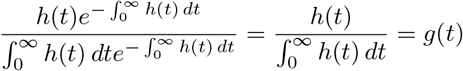

whereas if we require that the expected number of infections remains equal to *R*_*L*_, then 𝔼[*ν*] = *R*_*L*_.

The probability that no infections happen after transfer is then the sum of the probability of two independent events, that the single infection event happens prior to transfer, and that the event happens post-transfer, but results in no successful infections:

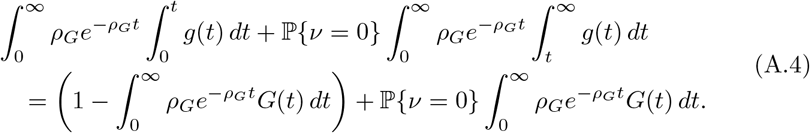

If, for example, we assume that *ν* is negative binomially distributed, say *ν* ∼ NB(*p, k*), then for a given *k*, we have 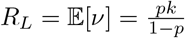, so 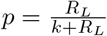 and

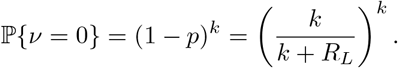

Finally, we note that

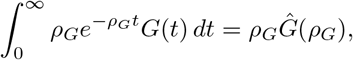

where *Ĝ*(*s*) is the Laplace transform of *G*(*t*). In particular,

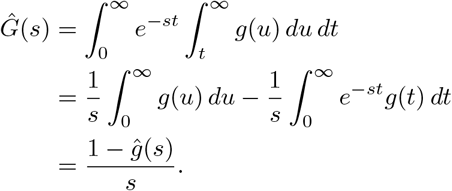

Thus, if we know *a priori* the distribution of the generation interval (*e.g*. lognormal, gamma or Weibull distributions are frequently posited), then the Laplace transform *ĝ*(*s*) = *M*_*g*_(−*s*), where *M*_*g*_(*s*) is the moment generating function is commonly available. Additionally, when *ρ*_*G*_ is small, we have

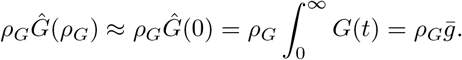

where 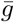 is the mean length of the generation interval.

### Appendix A.2. Probability of a Large Local Outbreak

#### Appendix A.2.1. Poisson Distributed Infections

To compute the probability of a local large outbreak under our generation interval approach, we must take care to distinguish between the first individual initiating the epidemic, for whom some fraction of the infectious period has already elapsed, and the newly infected individuals in the new site.

Under the assumption of a large local population, we can continue to use branching process recursive formulas to determine the probability that no locally infected individual gives rise to a significant outbreak, say *q*_*L*_:

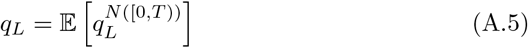

where *N* ([0, *T*)) is the number of individuals infected by a given individual before they are transferred at time *T* ; as before, *T* is exponentially distributed with rate *ρ*_*G*_, whereas conditional on *T, N* ([0, *T*)) is Poisson distributed with rate 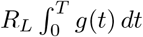. Now,

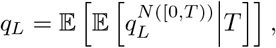

where the outer expectation is for the random variable *T*. Recognizing the inner expectation as the probability generating function for a Poisson random variable gives us

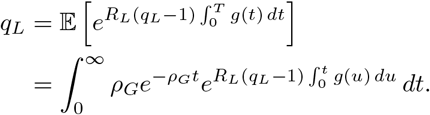

Now consider the initial infected individual. Let *T*, still exponentially distributed with rate *ρ*_*G*_ be the time after the start of their infectious period at which that individual was introduced into the local community. Then, they will infect *N* ([*T*, ∞)) individuals in the new site, whence the probability that there is not a major outbreak is

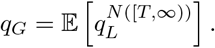

Again, first taking the conditional expectation of *N* ([*T*, ∞)) given *T*, which is a Poisson random variable with rate 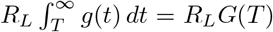, and then over *T*, gives

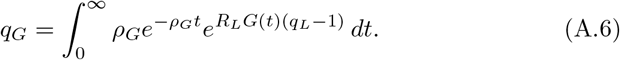

##### Appendix A.2.2. Individually Varying Reproductive Number

More generally, as before, we can consider the possibility that each individual has an i.i.d. reproductive number, say *R*_i_ drawn according to a Gamma 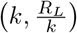 distribution, in which case, we may proceed as before to obtain (A.5), which we must now average over the gamma distribution to obtain

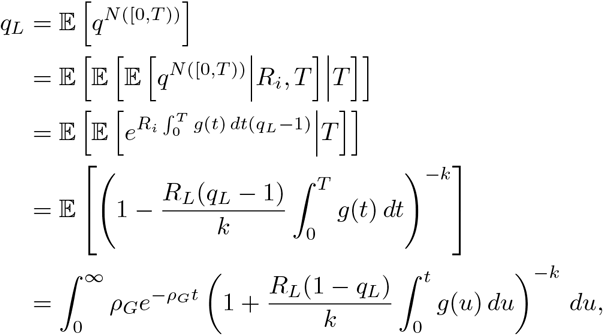

which we may solve numerically for *q*_*L*_. Proceeding similarly, averaging (A.6) over the gamma distribution and interchanging the order of integration yields

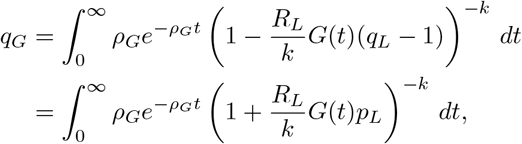

where *p*_*L*_ = 1 − *q*_*L*_.

#### Appendix A.2.3. Compound Poisson Processes

Now, suppose that at the *i*^th^ time of the inhomogeneous Poisson processes *N* ([0, *T*)) and *N* ([*T*, ∞)) (as previously, *T* indicates either the transfer time out of, or into, the focal site respectively) the individual independently infects *ν*_*i*_ individuals, where *ν*_*i*_ is a random variable with probability generating function *P*_*ν*_ (*z*) = 𝔼[*z*^*ν*^].

Now, since the *ν*_*i*_ are independently and identically distributed and independent of their arrival times in *N* ([0, *T*)),

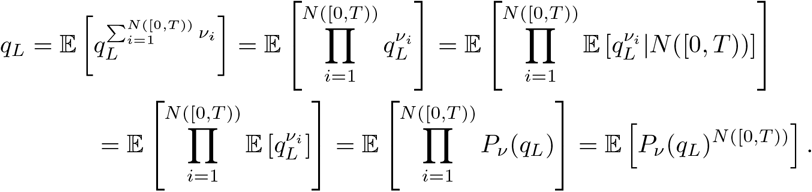

Now, proceeding as before,

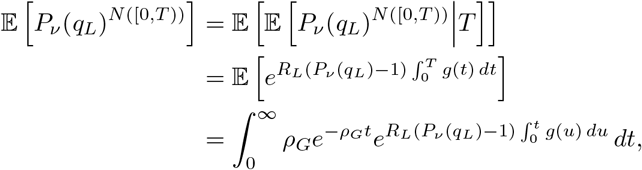

giving us the relation 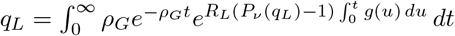.

Proceeding similarly gives us

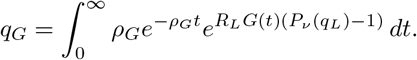

#### Appendix A.2.4. Bursts of Infections

Suppose each individual waits a randomly distributed time with probability density function *g*(*t*) before causing a random number *ν* of infections, independent of the time of transmission. Fix a given individual, and suppose that they are transferred at time *T*, exponentially distributed with rate *ρ*_*G*_ and that their transmission event happens at time *T′*. Then, their local chain of infection goes extinct if either they are transferred prior to transmission, or if not, if all those they infect have finite chains of infection:

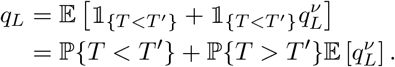

Again, the expectation on the right is the probability generating function for *ν* evaluated at *q*_*L*_, *P*_*ν*_ (*q*_*L*_).

The calculation thus depends on the choice of law for the random variables *ν*_*i*_, or equivalently the choice of distribution and its probability generating function *P*_*ν*_ (*z*). For example, if each *ν*_*i*_ is negatively binomially distributed with mean *R*_*L*_ and *k* successes (and thus success probability 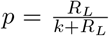) we have 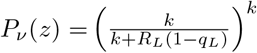, whereas for a Poisson process with the same mean *R*_*L*_ we have 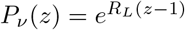.

On the other hand,

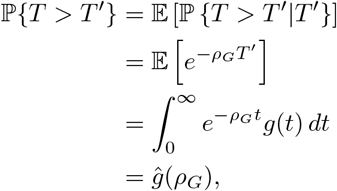

where *ĝ*(*p*) indicates the Laplace transform of *g*(*t*). Thus,

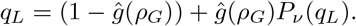

For the initial individual, we need to take into account the possibility that the individual had their “burst” prior to being transferred to the focal site,

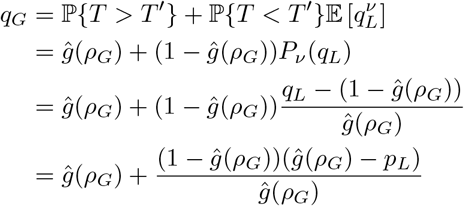

or, rearranging,

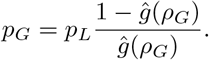

### Appendix A.3. Threshold mixing rates

Synthesizing the above results to derive a threshold rate of transfers, we recall that the criterion for a major outbreak on the congregate level is *R*_*∗*_ > 1, where *R*_*∗*_ = *µ* 𝔼[*Z*], *µ* is the (size-biased) mean size of a major outbreak, and *Z* is the number of facilities in which a given individual causes a major outbreak. Assuming that each individual is transferred at most once, then 𝔼[*Z*] = *p*_*G*_ = 1 − *q*_*G*_, where *q*_*G*_ is calculated as above. To calculate the critical transfer rate 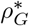, one needs to solve *R*_*∗*_ = 1 for *ρ*_*G*_.

## Appendix B. Approximating the Final Size of the Epidemic

Given the large populations in individual prisons, we approximate the proportion of susceptible, exposed and infective individuals by the classical SIR ordinary differential equations [11]:

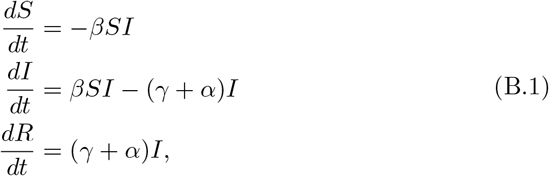

where *β* is the individual contact rate, whereas *γ* is the rate of recovery and *α* is the rate of disease-induced mortality. (For the SIR model and a wide class of related models, the error in making this deterministic approximation to the frequencies in each class in a population of *N* individuals is order 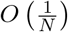; see [12] for details).

Whilst these equations don’t admit an analytic solution, one can solve for the trajectory in phase space, as Kermack & McKendrick observed in their landmark paper. Dividing the first equation in (B.1) by the third yields

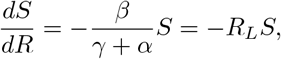

Whence 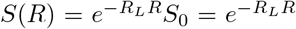, assuming the entire population is initially susceptible. Let *S*_*∞*_ and *R*_*∞*_ be the fraction susceptible and recovered at the end of the epidemic. By definition, the epidemic ends when no infected individuals remain, whence

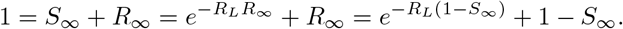

Rearranging yields 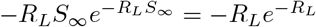. This equation may be solved via Lambert’s *W* -function [6], the transcendental (multi-)function satisfying *x* = *W* (*x*)*e*^*W* (*x*)^. In particular,

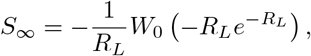

where *W*_0_(*x*) is the principal branch of the *W* function, real valued and increasing on [—*e*^*−*1^, ∞), so *S*_*∞*_ is a decreasing function of *R*_*L*_.

Moreover, *W*_0_(*x*) = *x* + *O*(*x*^2^), and bounded below by x, so that for large values of 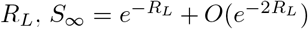. For *R*_*L*_ = 4, we have that 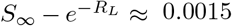, whereas *S*_*∞*_ ≈ 0.0198, so the relative error in approximating *S*_*∞*_ by 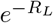 is already less than 8%, whereas approximately 98% of the population will have been infected, thus justifying the use of the mean household size as a reasonable upper bound on the final size of the epidemic.

We further note that for all values of 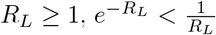 and the remaining fraction susceptible is well below the threshold of so-called herd immunity (the latter indicates the point at which epidemic growth decelerates, not the end of the epidemic).

In particular, after a first epidemic has run its course, assuming no change in the fraction susceptible, the total number infected due to a subsequent global infection is bounded above by a *subcritical* branching process with per-capita birth and death rates *βS*_*∞*_ and *γ* + *α* respectively. The expected total progeny of this branching process is 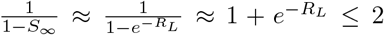. Thus, when households are very large, the impact of subsequent importations of infection have a negligible effect on the final fraction infected.

We remark that the argument above is unchanged by introducing a latent period of average length *θ* and a class of exposed, but non-infective individuals (the SEIR model):

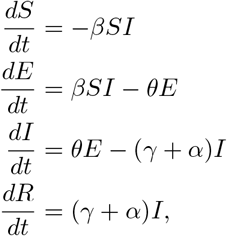

so the expression (and approximations) derived for *S*_*∞*_ remain valid (we note that in the absence of natural mortality, 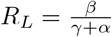 for the SEIR model as well).

In modelling COVID-19, it is also important to include an asymptomatic class that transmits the virus but does not experience excess mortality. We assume that a fraction *p* of all exposed individuals become asymptomatic spreaders, who can either recover or subsequently enter the symptomatic class at rate *ν*:

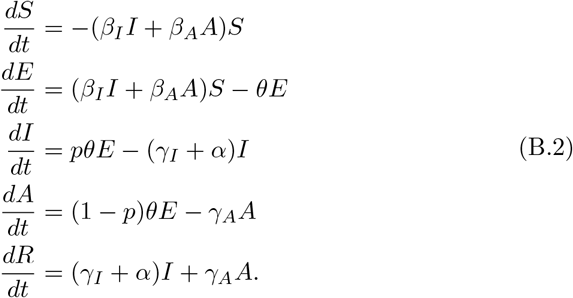

While this model is no longer analytically tractable, even in phase space, we can still obtain upper and lower bounds for the final size of the epidemic. Set 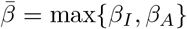 and <u>*β*</u> = min{*β*_*I*_, *β*_*A*_} and define 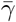 and <u>*γ*</u> similarly. Then,

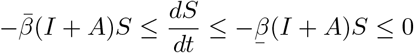

and

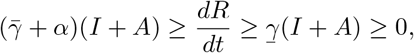

so that

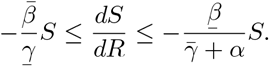

The comparison principle then tells us that

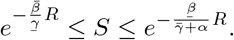

Proceeding as before, we find that

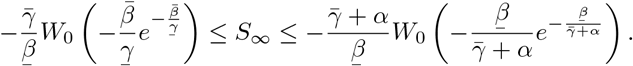

We can compute *R*_*L*_ using the next generation matrix method of [7]: linearized about the initial state with all individuals susceptible, the dynamics of the infected classes are described by a matrix equation:

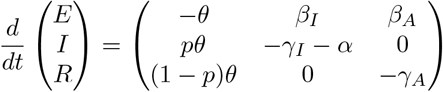

The transition matrix above is decomposed into matrices of *transmissions* (rates of new infections) and *transitions* (rates of events by which infected individuals change state):

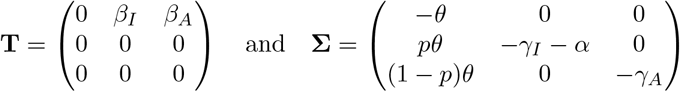

respectively. The *next generation matrix* is then

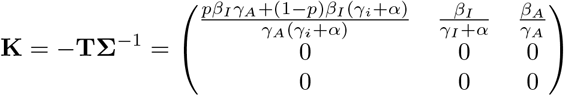

Theorem A.1 in [7] then tells us that 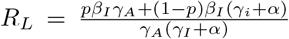. Thus, the threshold for herd immunity must lie in the range 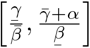.

## Footnotes

1 https://www.themarshallproject.org/2020/12/18/1-in-5-prisoners-in-the-u-s-has-had-covid-19, retrieved January 29, 2021.

2 https://www.nytimes.com/interactive/2020/us/coronavirus-us-cases.html, retrieved January 29, 2021.

3 UCLA Covid Behind Bars Project, retrieved March 26, 2021 from https://github.com/uclalawcovid19behindbars/data.git

4 https://www.courts.ca.gov/opinions/documents/A160122.PDF, retrieved January 29, 2021.

5 The Amend Project, *Urgent Memo COVID-19 Outbreak: San Quentin Prison*, retrieved January 29, 2021 from https://amend.us/wp-content/uploads/2020/06/COVID19-Outbreak-SQ-Prison-6.15.2020.pdf.

6 Retrieved March 26, 2021 from https://github.com/uclalawcovid19behindbars/data.git.

7 Population sizes published by CDCR, retrieved April 24, 2021 from https://www.cdcr.ca.gov/research/wp-content/uploads/sites/174/2021/03/Tpop1d210324.pdf, California City Correctional Facility population retrieved April 24, 2021 from https://www.cdcr.ca.gov/covid19/population-status-tracking/.

8 https://www.cdcr.ca.gov/covid19/wp-content/uploads/sites/197/2021/01/COVID-19-Screening-and-Testing-Matrix-Final-21-01-08.pdf, accessed February 4, 2021.

